# The impact of residency training in family medicine on hospital admissions due to Ambulatory-care Sensitive Conditions in Rio de Janeiro

**DOI:** 10.1101/2022.07.01.22277166

**Authors:** Adelson Guaraci Jantsch, Bo Burström, Gunnar Nilsson, Antônio Ponce de Leon

## Abstract

**BACKGROUND:** Lack of skilled human resources in primary care remains a major concern for policymakers in low- and middle-income countries. There is little evidence supporting the impact of residency training in family medicine in the quality of care, and it perpetuates misconceptions among policymakers that the provision of primary care can be easily done by any physician without special training.

**METHODS:** This article compares the risk of patients being hospitalized due to Ambulatory care sensitive conditions and the odds of having follow-up visits in primary care after hospital discharge, according to the type of their medical provider: (1) Generalists (reference), (2) Family physicians; and, (3) patients with no consultations prior to the event. Multilevel multivariate binomial regression models estimated the relative risks of a patient being hospitalized in a given month and the relative risks for the occurrence of a follow-up visit in primary care in a retrospective cohort of 636.640 patients between January 2013 and July 2018 in Rio de Janeiro.

**FINDINGS:** For all 14 conditions, there was a higher risk of hospitalization when patients had no consultation in primary care prior to the event. Except for Ear, Nose and Throat infections, patients seen by family physicians had a lower risk of being hospitalized, compared to patients seen by Generalists. Follow-up visits were more likely to happen among patients treated by family physicians for almost every condition analyzed.

**CONCLUSION:** With two years of training in family medicine, Family physicians can reduce the risk of their patients being hospitalized and increase the likelihood of those patients having a follow-up consultation in primary care. Investments in residency training in family medicine should be made to fix the shortage of skilled physicians in primary care, reduce hospitalizations and improve quality and continuity of care.

## INTRODUCTION

Hospital admissions have been used over the last 30 years as a valuable indicator of the effectiveness of Primary Health Care (PHC).^1^ Health conditions for which good quality, timely and effective primary care can prevent hospitalization are known as ambulatory-care sensitive conditions (ACSC). Good quality PHC services can change the course of these health conditions, making a hospital admission less likely to happen.^2,3^ High rates of hospitalizations due to ACSC would represent the provision of suboptimal PHC to the community.

This concept has been used worldwide to measure the impact of PHC initiatives, policies, and services,^4–7^ and in Brazil to show the impact of the Family Health Strategy^8^ (FHS), a federal policy that organizes the public PHC system, providing the structure and financial resources for Family Health Teams (FHTs). The FHS today – with nearly 43 000 FHTs in place – is responsible for the provision of PHC for 64% of the Brazilian population.^9,10^ Over its 25 years of implementation, many achievements have been reported, such as the reduction of infant, neonatal,^11–13^ and cardiovascular deaths,^14,15^ and for the reduction of hospital admissions due to ACSC,^16^ and among older adults.^17,18^

However, the lack of trained human resources in PHC remains a major concern for policymakers in low- and middle-income countries. Recent policies – the More Doctors and the Doctors for Brazil programs – have tried to promote provision and fixation of physicians in underserved areas.^19,20^ Nonetheless, the vast majority of doctors working at the FHS have no training in family medicine (FM).^21^ Despite the recent incentives to boost residency training in family medicine (RTFM) in the country, with less than 4% of the total number of vacancies for residency training dedicated to FM,^22^ this picture is far away from being changed. Unfortunately, there is little of evidence showing the impact of RTFM on the quality of care, and it perpetuates misconceptions among policymakers that the provision of primary care can be easily done by any physician without RTFM.

This study tests the hypothesis that RTFM can make doctors more capable of providing good quality primary care and prevent the occurrence of hospitalizations due to ACSC. The rationale behind this argument is that during two years of RTFM, FPs would develop the skills and competencies needed to better manage a wider set of conditions in primary care, thus preventing unnecessary hospital admissions. This article aims to (1) compare the risk of patients being hospitalized due to ACSC and (2) the odds of a patient having follow-up visits in primary care after hospital discharge, according to the type of training of their medical health care provider. Finally, it (3) estimates the population attributable fraction and the absolute change in hospital admissions per year in the sample that would happen if all medical consultations were performed by specialists in family medicine.

## METHODS

### STUDY DESIGN AND DATA SOURCE

A retrospective longitudinal analysis of medical consultations in primary care and hospital admissions in the Rio de Janeiro municipality was performed using patients’ information from electronic health records and from the National Hospital Admissions System. Deterministic linkage combining patient’s name, date of birth, mother’s name, gender and patient’s address was performed and false-positive matches between the datasets were excluded after manual review. Patients’ consent was not necessary since only anonymized information was used during the study and the Rio de Janeiro Municipal Health Department, the actual caretaker of this information, gave the consent to use this dataset for this research. The study was approved by the Rio de Janeiro Municipal Health Department research ethics board and it is registered under the number 03795118.0.0000.5279. It was conducted in accordance with the 466/12 resolution from the Brazilian National Health Council and the Declaration of Helsinki.

### EXPOSURE AND INDEPENDENT VARIABLES

Physicians were categorized according to their type of formal training to work in PHC: (1) Generalists, i.e. doctors without RTFM, were considered as the reference category; and (2) Family physicians (FPs) – graduated FPs, FM preceptors and residents enrolled in the FM residency programs. Patients that were hospitalized but had no consultations in primary care prior to the event were categorized as (3) “No consultation prior”.

Residents and preceptors in FM share responsibilities for the same patients at the same FHT for two years of training and the decisions the residents make are usually supervised by their preceptor, hence they were both considered as FPs. Residents work 48 hours per week under the full supervision of FM preceptors in a community-based primary care clinic. They have learning sessions every week addressing topics of FM and PHC and clinical content about the most prevalent health conditions in primary care. All learning activities the residents must take during the course are designed in line with the National Committee for Medical Residencies and with the Brazilian Society of Family and Community Medicine (SBMFC).^23^ Information about other forms of post-graduate training or specialization was not available and were not taken into account, nor the number of years in practice for any doctor.

Patients contributed to the models with information about (1) age (continuous), (2) sex, and (3) the Charlson Comorbidity Index.^24^ Each patient contribute only from the date of subscription at the clinic until the last day available at the dataset, or the day the patient was unsubscribed or the date of death. The Social Development Index (SDI) was used to bring information about the context where the patient lives. The SDI is a linear scale combining information about sanitation, schooling, income, and housing conditions from every household in the FHT catchment area, representing the grade of social development of a neighborhood. Hence, patients registered in the same FHT have the same SDI. It varies from 0 (least developed) to 1 (most developed).

All clinics and FHT in this sample have the same physical structure, offices equipped with computer, printer, medical equipment, room for small surgical procedures, the same arsenal of laboratory tests and medicines in the pharmacy, and the same type of human resources available: nurses, technicians, dentists, pharmacists, and managers. The distribution of doctors among different clinics and FHT didn’t follow any criteria that is known to interfere and generate confounding in the relationship between the medical categories, the population assisted, and the study outcomes.

### OUTCOMES

Two outcomes were considered in this study: (1) The occurrence of hospital admissions, and (2) the occurrence of a follow-up visit in primary care in the following two, four, and six months after patient’s discharge from the hospital.

Fourteen health conditions from the Brazilian list of Ambulatory-care Sensitive Conditions (ICSAP-Brazil) were analyzed. ICD-10 codes and clinical criteria for each condition were used to identify both population-at-risk and the event, i.e. the hospital admissions – table 1. Both hospital admissions and follow up visits were considered as binary events, i.e. being or not being hospitalized in a given month among those at risk and having or not having one medical consultation within 60 days, 120 days, or 180 days after being discharged from the hospital. For the follow-up visits, patients that were hospitalized and have not died during hospitalization were considered as the population-at-risk.

**Table 1:**
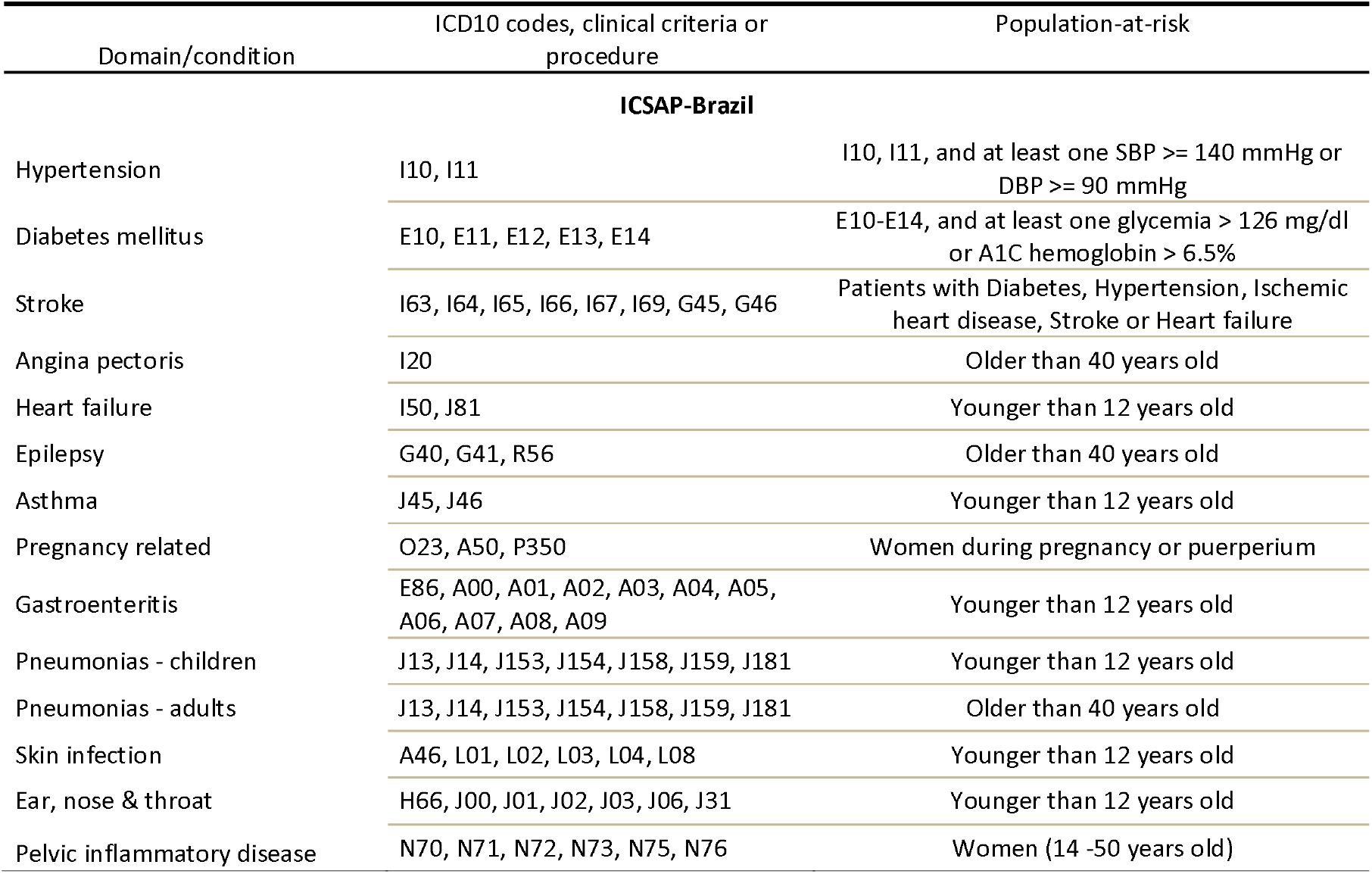
List of health conditions according to their ICD10 codes and clinical criteria and respective population-at-risk used in the regression models.

By studying both phenomena here we aim to capture both sides of coordination of care between primary and secondary/tertiary care. If RTFM can make doctors more competent to work in primary care, it will make hospital admissions less likely to occur and, at the same time, for those patients that have been hospitalized, it will make a follow-up visit more likely to occur.

### STATISTICAL ANALYSES

Multilevel multivariate binomial regression models estimated the relative risks (RR) of a patient being hospitalized in a given month and the RR for the occurrence of a follow-up visit, according to the type of doctor taking care of the patient, i.e. (1) Generalists (reference), (2) FPs, and, for those with no consultations prior to the event, (3) “No consultation prior”. A two levels hierarchical data structures was created clustering months within patients. This nested design was used in order to regard the data dependence.

For follow-up visits in primary care, only patients that were hospitalized and did not die during hospitalization were considered as population-at-risk. Time effects were regarded using dummy variables for months and years. All models were adjusted for patient information (sex, age, and Charlson comorbidity index) and contextual information (SDI and time). For both acute and chronic conditions, patients were considered “at risk” during the whole period they were subscribed at the clinic. For Pregnancy related hospital-admissions, only pregnant women and women during the puerperium were considered as patients-at-risk.

Finally, Population Attributable Fraction (PAF) for each outcome was calculated using the RR from the regression models to estimate the absolute change In terms of number of hospital admissions per year in the sample that would happen if all medical consultations were performed by trained FPs. Data processing and statistical analysis were performed using R version 3.6.2 and lme4 package.

## RESULTS

From the original list of 636.640 patients (376.204 women and 261.432 men), 457.533 (184.505 men and 273.662 women) were subscribed in 195 FHTs and 30 different PHC clinics. From the patients assigned to one FHT, 406.271 patients were responsible for 2.433.924 medical consultations between January 2013 and December 2018, and for 80.314 hospital admissions between January 2013 and July 2018 that occurred among 59.106 different patients.

For all 14 domains studied and for both hierarchical models, there was a higher risk of hospitalization when patients had no consultation in primary care prior to the event. This effect was higher when the cause of hospitalization was due to Stroke, Gastroenteritis and Pelvic inflammatory disease, and had its smallest value in cases of Asthma and Epilepsy.

Except for Ear, Nose and Throat infections, patients seen by FPs tended to have a lower risk of being hospitalized, compared to patients seen by Generalists. This effect was greater in hospital admissions due to Asthma, Pneumonias in adults, Gastroenteritis, Heart failure and Pneumonia in children.

In absolute numbers, Heart failure, Angina pectoris, Pneumonias in adults would express the biggest decrease in the total number of hospital admissions, with 68, 61 and 59 fewer hospitalizations every year, if all doctors working in FHTs were trained in FM residency programs (table 3). In this scenario, there could be a 47% reduction in hospital admissions for Asthma, a 50% reduction in Pneumonia in adults and a 31% reduction in Pelvic inflammatory diseases.

**Table 2:** Number of medical consultations and patients’ characteristics according to each medical category in the study sample. Rio de Janeiro, Brazil, 2015 – 2018.

| Medical category | Number of doctors – N (%) | Consultations – N (%) | SDI -mean (SD) | Patients' age (%) |  |  | Patients according to sex – N (%) |  |
| --- | --- | --- | --- | --- | --- | --- | --- | --- |
|  |  |  |  | < 18 | > 18 & < 45 | > 45 | Women | men |
| Generalists | 633 (75.6) | 1.629.235 (67.5) | 0.573 (0.03) | 21.3 | 31.8 | 46.9 | 1.067.212 (65.6) | 562.023 (34.4) |
| Family physicians | 204 (24.4) | 785.273 (32.5) | 0.585 (0.03) | 18.5 | 34.5 | 47.0 | 517.813 (65.9) | 267.460 (34.1) |

**Table 3:** Relative risks for hospital admissions from January 2013 to July 2018 according to the proportion of consultations performed by Family physicians or Generalists in a Family Health Team in Primary Care. Rio de Janeiro, Brazil. 2013-2018.

|  | Family physicians | No consultation prior |
| --- | --- | --- |
| Hypertension | 0.83 (0.61; 1.15) | 2.63 (2.04; 3.40) |
| Diabetes mellitus | 0.76 (0.56; 1.04) | 2.75 (2.11; 3.60) |
| Stroke | 0.74 (0.58; 0.95) | 5.47 (4.50; 6.65) |
| Angina pectoris | 0.62 (0.51; 0.75) | 2.58 (2.19; 3.03) |
| Heart failure | 0.52 (0.40; 0.67) | 2.78 (2.19; 3.53) |
| Epilepsy | 0.79 (0.57; 1.11) | 1.40 (1.07; 1.83) |
| Asthma | 0.35 (0.21; 0.57) | 1.46 (1.04; 2.05) |
| Pregnancy related | 0.78 (0.60; 1.01) | 2.89 (2.30; 3.63) |
| Gastroenteritis | 0.45 (0.25; 0.79) | 3.17 (2.19; 4.59) |
| Pneumonias - children | 0.57 (0.42; 0.77) | 2.19 (1.74; 2.76) |
| Pneumonias - adults | 0.36 (0.24; 0.56) | 2.90 (2.13; 3.95) |
| Skin infection | 0.82 (0.69; 0.97) | 2.61 (2.23; 3.00) |
| Ear, nose & throat | 1.06 (0.64; 1.77) | 1.78 (1.13; 2.83) |
| Pelvic inflammatory disease | 0.59 (0.38; 0.93) | 3.35 (2.34; 4.81) |

Multilevel multivariate binomial regression models adjusted for age, sex, time, and the Social development index of the area covered by the FHT.

For follow-up visits (table 4), patients that had no prior visit at the clinic before hospitalization were less likely than those seen by Generalists to have a follow-up consultation in primary care after hospital discharge. This effect is almost the same for consutations within two, four and six months. Oppositely, follow-up visits were more likely to happen among patients treated in FHTs with FPs for almost all conditions analyzed. Patients treated by FPs that were hospitalized for Heart failure and Angina pectoris had a consistent higher odds of having follow-up consultations. The exceptions were hospital admissions due to Pregnancy-related and Gastroenteritis, where the odds of a follow-up consultation happening were the same for patients seen by Generalists and FPs.

**Table 4:**
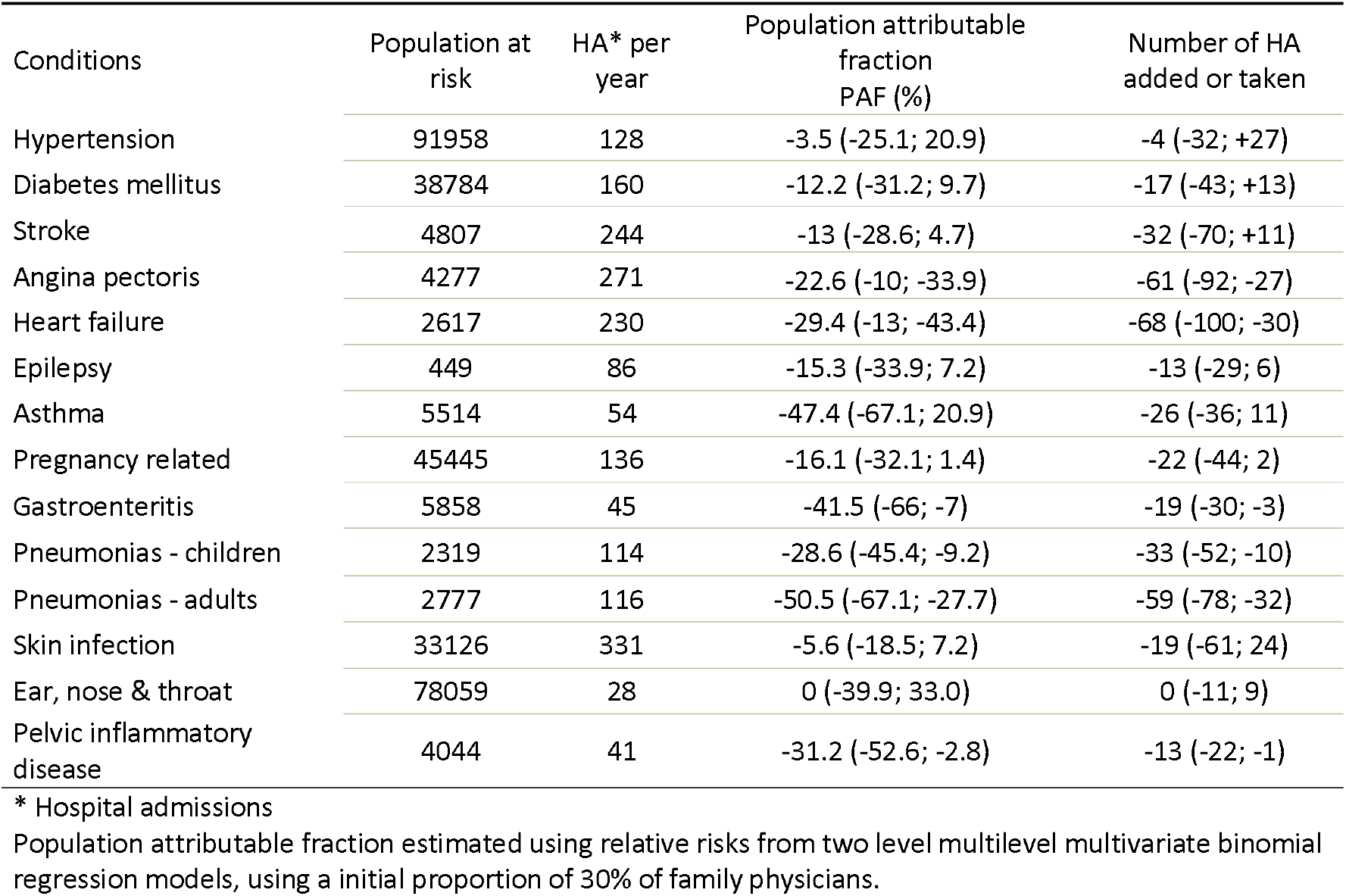
Population attributable fraction and the absolute change in the number of hospital admissions in 12 months in a scenario where all Family Health Teams in the sample would have trained Family physicians as medical professionals. Rio de Janeiro, Brazil, 2013-2018.

| Conditions | Population at risk | HA* per year | Population attributable fraction PAF (%) | Number of HA added or taken |
| --- | --- | --- | --- | --- |
| Hypertension | 91958 | 128 | -3.5 (-25.1; 20.9) | -4 (-32; +27) |
| Diabetes mellitus | 38784 | 160 | -12.2 (-31.2; 9.7) | -17 (-43; +13) |
| Stroke | 4807 | 244 | -13 (-28.6; 4.7) | -32 (-70; +11) |
| Angina pectoris | 4277 | 271 | -22.6 (-10; -33.9) | -61 (-92; -27) |
| Heart failure | 2617 | 230 | -29.4 (-13; -43.4) | -68 (-100; -30) |
| Epilepsy | 449 | 86 | -15.3 (-33.9; 7.2) | -13 (-29; 6) |
| Asthma | 5514 | 54 | -47.4 (-67.1; 20.9) | -26 (-36; 11) |
| Pregnancy related | 45445 | 136 | -16.1 (-32.1; 1.4) | -22 (-44; 2) |
| Gastroenteritis | 5858 | 45 | -41.5 (-66; -7) | -19 (-30; -3) |
| Pneumonias - children | 2319 | 114 | -28.6 (-45.4; -9.2) | -33 (-52; -10) |
| Pneumonias - adults | 2777 | 116 | -50.5 (-67.1; -27.7) | -59 (-78; -32) |
| Skin infection | 33126 | 331 | -5.6 (-18.5; 7.2) | -19 (-61; 24) |
| Ear, nose & throat | 78059 | 28 | 0 (-39.9; 33.0) | 0 (-11; 9) |
| Pelvic inflammatory disease | 4044 | 41 | -31.2 (-52.6; -2.8) | -13 (-22; -1) |

\* Hospital admissions
Population attributable fraction estimated using relative risks from two level multilevel multivariate binomial regression models, using a initial proportion of 30% of family physicians.

**Table 5:**
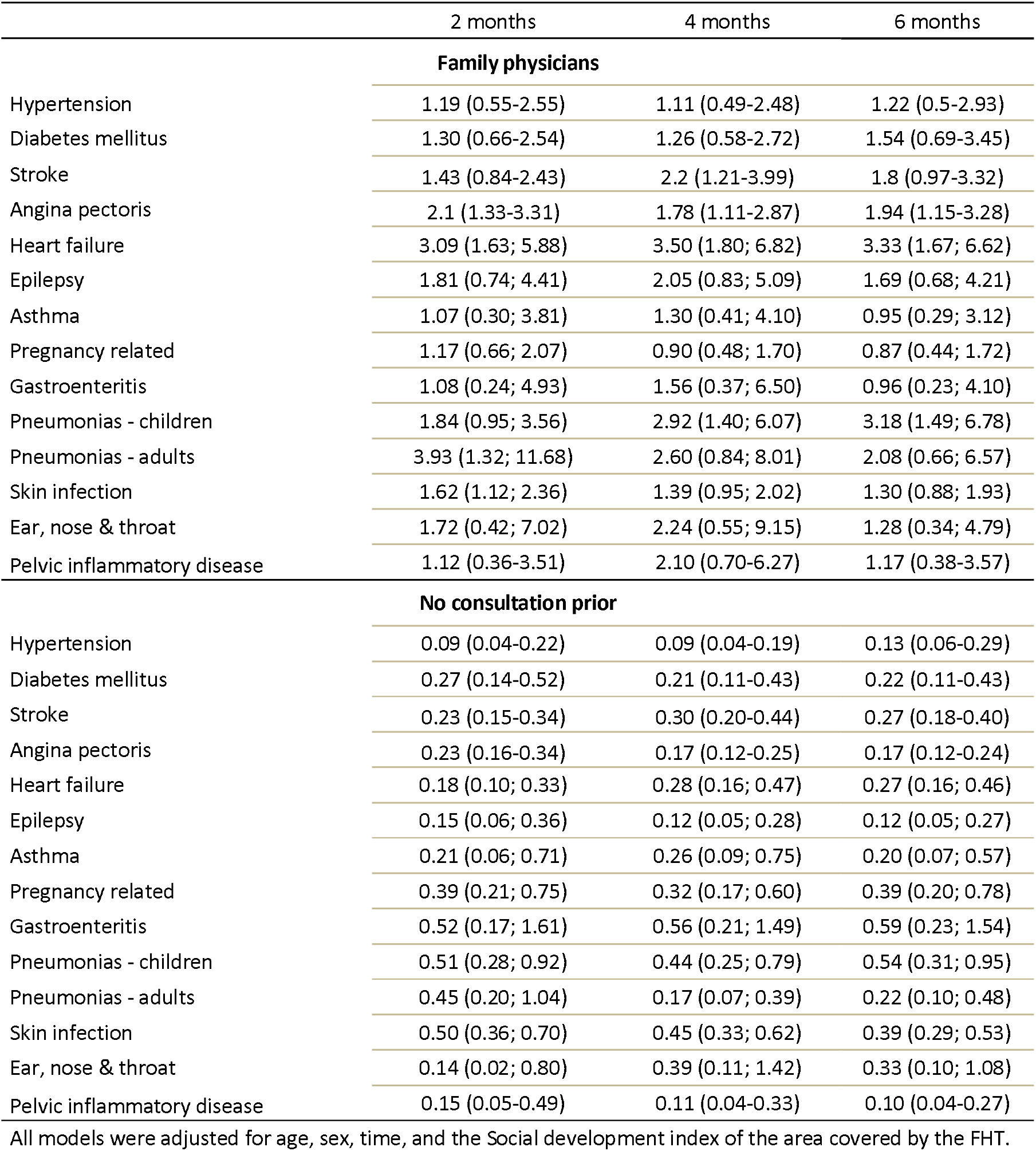
Relative risks for a patient having a follow-up consultation in primary care after one, three, and six months after hospital discharge according to the proportion of medical consultations performed by Family physicians or Generalists in a Family Health Team in the previous twelve months. Rio de Janeiro, Brazil.

|  | 2 months | 4 months | 6 months |
| --- | --- | --- | --- |
| **Family physicians** |  |  |  |
| Hypertension | 1.19 (0.55-2.55) | 1.11 (0.49-2.48) | 1.22 (0.5-2.93) |
| Diabetes mellitus | 1.30 (0.66-2.54) | 1.26 (0.58-2.72) | 1.54 (0.69-3.45) |
| Stroke | 1.43 (0.84-2.43) | 2.2 (1.21-3.99) | 1.8 (0.97-3.32) |
| Angina pectoris | 2.1 (1.33-3.31) | 1.78 (1.11-2.87) | 1.94 (1.15-3.28) |
| Heart failure | 3.09 (1.63; 5.88) | 3.50 (1.80; 6.82) | 3.33 (1.67; 6.62) |
| Epilepsy | 1.81 (0.74; 4.41) | 2.05 (0.83; 5.09) | 1.69 (0.68; 4.21) |
| Asthma | 1.07 (0.30; 3.81) | 1.30 (0.41; 4.10) | 0.95 (0.29; 3.12) |
| Pregnancy related | 1.17 (0.66; 2.07) | 0.90 (0.48; 1.70) | 0.87 (0.44; 1.72) |
| Gastroenteritis | 1.08 (0.24; 4.93) | 1.56 (0.37; 6.50) | 0.96 (0.23; 4.10) |
| Pneumonias - children | 1.84 (0.95; 3.56) | 2.92 (1.40; 6.07) | 3.18 (1.49; 6.78) |
| Pneumonias - adults | 3.93 (1.32; 11.68) | 2.60 (0.84; 8.01) | 2.08 (0.66; 6.57) |
| Skin infection | 1.62 (1.12; 2.36) | 1.39 (0.95; 2.02) | 1.30 (0.88; 1.93) |
| Ear, nose & throat | 1.72 (0.42; 7.02) | 2.24 (0.55; 9.15) | 1.28 (0.34; 4.79) |
| Pelvic inflammatory disease | 1.12 (0.36-3.51) | 2.10 (0.70-6.27) | 1.17 (0.38-3.57) |
| **No consultation prior** |  |  |  |
| Hypertension | 0.09 (0.04-0.22) | 0.09 (0.04-0.19) | 0.13 (0.06-0.29) |
| Diabetes mellitus | 0.27 (0.14-0.52) | 0.21 (0.11-0.43) | 0.22 (0.11-0.43) |
| Stroke | 0.23 (0.15-0.34) | 0.30 (0.20-0.44) | 0.27 (0.18-0.40) |
| Angina pectoris | 0.23 (0.16-0.34) | 0.17 (0.12-0.25) | 0.17 (0.12-0.24) |
| Heart failure | 0.18 (0.10; 0.33) | 0.28 (0.16; 0.47) | 0.27 (0.16; 0.46) |
| Epilepsy | 0.15 (0.06; 0.36) | 0.12 (0.05; 0.28) | 0.12 (0.05; 0.27) |
| Asthma | 0.21 (0.06; 0.71) | 0.26 (0.09; 0.75) | 0.20 (0.07; 0.57) |
| Pregnancy related | 0.39 (0.21; 0.75) | 0.32 (0.17; 0.60) | 0.39 (0.20; 0.78) |
| Gastroenteritis | 0.52 (0.17; 1.61) | 0.56 (0.21; 1.49) | 0.59 (0.23; 1.54) |
| Pneumonias - children | 0.51 (0.28; 0.92) | 0.44 (0.25; 0.79) | 0.54 (0.31; 0.95) |
| Pneumonias - adults | 0.45 (0.20; 1.04) | 0.17 (0.07; 0.39) | 0.22 (0.10; 0.48) |
| Skin infection | 0.50 (0.36; 0.70) | 0.45 (0.33; 0.62) | 0.39 (0.29; 0.53) |
| Ear, nose & throat | 0.14 (0.02; 0.80) | 0.39 (0.11; 1.42) | 0.33 (0.10; 1.08) |
| Pelvic inflammatory disease | 0.15 (0.05-0.49) | 0.11 (0.04-0.33) | 0.10 (0.04-0.27) |

All models were adjusted for age, sex, time, and the Social development index of the area covered by the FHT.

## DISCUSSION

By way of clarification, the comparisons made here between Generalists and FPs are not judgmental and we believe that all doctors in this sample were doing their best to help their patients. We aimed solely to test the hypothesis that the addition of two years of training in family medicine, on top of the current six years of medical school in Brazil, modifies the risk of patients being hospitalized.

### First aim – impact of RTFM on the risk of hospital admissions due to ACSC

With two years of training in family medicine, Family physicians working in primary care can reduce the risk of their patients being hospitalized for all fourteen health conditions from the Brazilian list of Ambulatory-care Sensitive Conditions (ICSAP-Brazil), except for Ear, nose and throat infections. Moreover, patients subscribed to a FHT that had no consultations at the clinic were at a higher risk of been hospitalized for all conditions, compared to patients treated by Generalists or Family physicians. These findings add important information to the well-established notion that the increased coverage of FHS in Brazilian municipalities leads to lower hospital admissions,^5,16^ lower mortality from heart and cerebrovascular diseases,^14^ and lower mortality from ameanable conditions.^15^ These previous ecological studies, however, focused mainly on the number of FHTs available to the population, which translates to “the proportion of the population covered or not by a FHT in the municipality”. In other words, the higher the proportion of the population covered by the FHS in a municipality, the larger will be the impact on health outcomes. For the patients, being covered by a FHT – i.e., having one doctor, one nurse, one nurse-technician and community-health agents as a team of healthcare providers – makes the risk of a comprehensive list of events decrease, such as hospital admissions. What is new from our study is that the already known beneficial effect of being covered by a FHT increases if the doctor in charge is a Family physician. It also expands the evidence raised in previous studies showing that specialists in family medicine are more capable of detecting chronic health conditions, requesting fewer laboratory tests and fewer referrals to secondary care and providing more follow-up consultations to their patients in primary care.^25,26^

### Exploring the specific causes – infectious diseases

Exploring the specific causes for hospital admissions, with the exception of Ear, nose & throat infections and – to a lesser extent – Skin infections, all infectious disesases had their risk decreased when a Family physician was present at the FHT. Pelvic inflammatory disease, Gastroenteritis, Pneumonia (children and adults) and Gastroenteritis were strongly affected by the presence of a FP at the FHT and in the case of Pneumonias in adults more than 50% of all cases could be avoided if all FHT in this samples had trained family physicians as their healthcare provider. In all these acute conditions, with the exception of Gastroenteritis, it is possible that the treatment will require the use of antibiotics, which is a matter of concern in a scenario where there is no shortage of these drugs and health professionals can freely prescribe them. The prescription of antibiotics in ambulatory care is the subject of campaigns aiming to protect patients from unnecessary treatments and procedures. Choosing wisely is one of them and the rational use of antibiotics is a key competence that residents in FM need to develop.^27^ Future studies will need to explore the previous use of antibiotics for these patients-at-risk. Even if there is no difference in the risk of Hospital admission, like in Ear, nose & throat infections, if a lower prescription of antibiotics is found, it can represent a gain in terms of good practice.

Hospital admissions due to Gastroenteritis (diarrhea and dehydration) would also be influenced by RTFM, with a lower risk (RR 0.49 – 95%CI 0.26; 0.90) favoring patients treated by FPs. This lower risk is very unexpected, since this is an acute condition easily treated by a cheap and widely available intervention (oral rehydration salts). Moreover, Gastroenteritis is not a condition that challenges doctors to identify and treat, like heart failure, HIV, or dementia. Initiatives like the Integrated Management of Childhood Illness initiative – AIDIPI in Brazil – have been tackling this issue for many years by task shifting a simple procedure that, most of the time, does not require medical care. Nevertheless, biomedical content learned during RTFM cannot be entirely responsible for the lower risk of hospital admissions due to Gastroenteritis. Developing skills and attitudes that are beyond biomedical content represents a paradigm shift for medical education and a key element present in many competency-based curriculum in family medicine around the world.^23,28,29^

One can speculate whether the development of relational, communication and time management skills could increase patients’ trust in their doctors and change the way parents seek medical care for their sick children. The lower risk of Hospital admissions due to Asthma and Epilepsy for Family physicians can maybe be the consequence of the combination of these skills – clinical skills and competencies of family physicians. If patients with Asthma or Epilepsy in the case of an acute event have access to a competent physician able to manage the acute situation, most of the time it will be solved in the primary care setting. Looking for an answer to this question would require studies designed to explore the interaction of these skills and competencies and their effect on healthcare. This is not a simple research question to be pursued but it could help to highlight the pathways in which RTFM makes this happen.

### Exploring the specific causes – chronic conditions

Hypertension and Diabetes mellitus – Chronic cardiovascular conditions that play a major role as risk factors for cardiovascular outcomes – seemed to be less influenced by the presence of a FP in the FHT, when compared to Angina pectoris and Heart failure – outcomes that usually have Hypertension and Diabetes mellitus as their main causes. To a lesser degree, hospital admissions due to Stroke also seemed to be affected by the presence of FP in the FHT. There is a contradiction in these findings and it is reasonable to supose that treating Hypertension and Diabetes mellitus would decrease in a higher extent the risk for hospital admissions due to these conditions, and not so much the risk for their final consequences.

Lowering diabetic patients’ blood glucose reduces the risk of major cardiovascular events, especially for those patients with cardiovascular disease at baseline.^30^ Reducing systolic blood pressure significantly reduces coronary heart disease, stroke, and heart failure, with the effect increasing in that order.^31^ However, these measures would affect primarily Hypertension and Diabetes melittus, and a larger effect in the reduction of hospital admissions should be expected. However, different population-at-risk were considered for each outcome. For Hypertension, all patients with blood pressure higher than 140/90 mmHg were included as patients-at-risk of being hospitalized – the same procedure was undertaken for every condition. Hence, the population-at-risk for Diabetes mellitus and Hypertension ended up including patients with different health statuses – from mild cases that require no more than lifestyle changes and one diuretic pill, to more severe cases with multimorbidity and polypharmacy involved, with an extensive list of medicines, insulin regimens, and a higher morbidity burden. On the other hand, all patients-at-risk for Angina pectoris, Stroke and Heart failure had been diagnosed (at the hospital or at the primary care clinic) for these conditions, making all of them high-risk patients. The differences in terms of their health status are smaller, hence the population-at-risk for these conditions are more homogenous than that for Hypertension and Diabetes mellitus.

### Second aim – risk of patients having follow-up consultations in primary care after hospital discharge

Information on follow-ups also highlights an important aspect for the study of PHC and hospital admissions, which is continuity of care. Not only the risk of hospitalization was lower among patients treated by Family physicians but, for most conditions in our list, RTFM makes follow-up visits after hospital discharge more likely to happen. Since health care do not cease after hospital discharge, it seems that RTFM is also important in promoting continuity and coordination of care, making medical consultations more likely to happen after hospitalization. When compared to patients that had no consultation prior to the hospital admission (covered but not using the FHS), patients using their FHT as healthcare providers were more likely to have follow-up consultations. It is hard to tell what are the reasons for these patients to have fewer follow-up visits at the PHC clinic after hospital discharge. The same can be said about the reasons for these patients not to use their FHT before the hospital admission. Personal preferences for another healthcare service (private out-of-pocket or private health insurance), poor selfcare, and barriers to access the public primary care clinic are some possible reasons that should be explored in future studies.

### Third aim – absolute reduction in hospital admissions

Reporting the Population attributable fraction and the total number of hospital admissions avoided in a year can better translate the information reported as relative risks into meaningful information for policymakers and healthcare managers. In table 4 the relative risks are expressed in terms of the absolute number of hospital admissions that would happen if all FHTs had a family medicine specialist as medical provider. Some conditions would experience a big relative change with a small absolute reduction in the number of hospital admissions. This is the case for Asthma and Pelvic inflammatory disease. Even with 47% and 31 % fewer hospital admissions every year, the total number of events avoided would be only 26 and 13, respectively. Meanwhile, Angina pectoris and Heart failure – conditions with a larger number of hospital admissions per year – would experience a large absolute reduction of events (61 and 68, respectively), even with a lower relative risk reduction between family physicians and generalists.

Every avoided hospitalization due to ambulatory care sensitive conditions represents healtcare resources that are saved to better use with patients in need. Moreover, health care conditions will be differently affected by RTFM (relative risks) but will also result in different number of events, with different costs and different time number of days in the hospital. This dimension of the impact that RTFM can make in patient care was not measured in our study but would be an interesting focus for future research exploring duration of stay, quality of life and outcomes related to the very hospital admission.

### Methodological considerations and implications for future research

Exploring in detail the ways that Family physicians interact with their patients to change the life course of these conditions was not the aim of this study. Future research describing more precisely these populations-at-risk would be extremely valuable to explore the actions taken by family physicians and their patients during their health care trajectory that are responsible for lowering the risk for hospital admissions. Furthermore, each health condition in each patient might have a different time lapse to change under the effect of having a Family physician as his/her medical professional. Prenatal care issues and other acute conditions are more likely to be affected in a short term. A urinary tract infection in a pregnant woman develops over the period of one month (from the first symptoms to its resolution). On the contrary, chronic conditions resulting from years of exposure to risk factors will lead patients to a series of appointments, blood tests, follow-ups, changes in drug treatments.

In the case of Pregnancy related conditions, the lower risk of a women being hospitalized when a Family physician is responsible for her prenatal care is not followed by a change in the risk of having follow-up visits after hospital discharge. This information shows us that, in this sample, there were doctors available in the FHT to see these women after hospital discharge. Probably there was a doctor in that FHT before a hospital admission has occurred as well.

Hospital admissions avoided and follow-up visits after hospital discharge are only the tip of the iceberg. FPs can prevent these hospital admissions on top of changing a series of events that happen every day, such as the detection of health conditions, follow up consultations, procedures, prescriptions, and referrals to secondary care.

The ICSAP-Brazil creates a comprehensive list of conditions to explore and measure the effectiveness of PHC services, but it keeps out of its framework conditions that are widely prevalent in the community, such as Mental health conditions, HIV/AIDS, and Cancer. Those are some examples of prevalent conditions that might have the risk of hospitalization affected by strong PHC services and, moreover, by skilled Family physicians. Expanding the scope of health care conditions would be necessary to further explore in which extent RTFM affects the occurrence of hospital admissions.

### Implications for healthcare

The World Health Organization in 2020 published the Operational Framework for Primary Health Care^32^ describing the health care workforce as one of the 14 strategic levers to enable the development of PHC. Since the inauguration of the FHS in 1994,^8^ some of the 14 levers were prioritized: an organized model of care was established for the whole country, strengthening governance and social participation, and creating policy frameworks organizing the way PHC should be funded, delivered and how resources should be allocated. As mentioned before, the More Doctors and the Doctors for Brazil programs are good examples of policies that have tried to move the Healthcare workforce lever up, promoting provision and fixation of physicians in underserved areas. This is specially important when we look at the relative risks for Hospital admissions between patients with no consultation prior compared to those seen a Generalist. In a city with one of the highest density of doctors in the country,^22^ having a Family physician as a medical healthcare provider is beneficial to decrease the number of Hospital admissions but having consultations with a physician and using the FHT is crucial. For the rest of the country and especially in remotes areas, it is essential that the federal government fixes this inequality by providing physicians for places that can hardly get any.

Nevertheless, one more step must be taken by the federal government to leverage this lever, which is to fix the unequal distribution of vacancies for residency training in the country by increasing the number of seats for family medicine and redistributing these seats accross the country. The political structure for these programs is already in place and the financial resources are available. In other words, the country already has the foundation to build a national program for provision, fixation and training of physicians in primary care aligned with the established family medicine residency programs.

Only with investments in RTFM it will be possible to fix the shortage of skilled physicians we see today in Brazilian PHC.^21,22^ To make it happen, we need to correct common misconceptions among policymakers that doctors do not need residency training to work in primary care.

### STRENGTHS AND LIMITATIONS

The main limitation of our study is the lack of information regarding the usage of emergency care services in the city to fully cover all paths between primary care and hospital care. These services play an important role in managing every domain at the ICSAP-Brazil list, especially acute conditions and exacerbations of chronic diseases. Certainly, these services can avoid hospitalizations by providing short term treatment, and discharging patients to complete their treatment at home. Another limitation is the lack of information about private health care services in the community. Having the complete information would help to clarify if patients were sent to the hospital by the public primary care clinic, by a private clinic or by the emergency service, providing better information to clarify if there was a lack of access to the FHTs.

Relative risks, most of the time, cannot reach a broad audience. By reporting the population attributable fraction, we tried to translate relative risks into absolute numbers to provide more palatable information for decision makers and politicians get better understanding of the impact of RTFM. The total number of hospital admissions that would be added or taken if all FHTs had trained Family physicians as their medical provider can more easily reach those responsible for deciding to implement an educational initiative for capacity building of human resources for primary care.

Finaly, using real-world data to analyze an actual intervention in a middle-income country may reverberate better to other countries that are also trying to expand and improve their PHC systems.

## CONCLUSION

The Brazilian Family Health Strategy has been responsible for an impressive impact on public health for the last 25 years, decreasing health inequalities by strengthening governance and providing structure and financial support for FHTs in the whole country. Only recently, capacity building of human resources for primary care – especially residency training in family medicine – has been considered as an important piece for the success of this national policy. In this study, we presented evidence that public investments in capacity building of human resources for primary care can prevent the occurrence of hospital admissions.

It will take a lot of effort from all stakeholders and political will to allocate financial resources today in educational initiatives that will take some years to pay back in health outcomes the investment made. In the case of the medical workforce for primary care – especially in LMIC – this will not be achieved without investments in residency training in family medicine.

## Data Availability

Patients consent was not necessary since only anonymized information was used during the study and the Rio de Janeiro Municipal Health Department, the actual caretaker of this information, gave the consent to use this dataset for this research.

https://www.rio.rj.gov.br/web/sms/comite-de-etica-em-pesquisa

